# The accuracy of polygenic score models for anthropometric traits and Type II Diabetes in the Native Hawaiian Population

**DOI:** 10.1101/2023.12.25.23300499

**Authors:** Ying-Chu Lo, Tsz Fung Chan, Soyoung Jeon, Gertraud Maskarinec, Kekoa Taparra, Nathan Nakatsuka, Mingrui Yu, Chia-Yen Chen, Yen-Feng Lin, Lynne R. Wilkens, Loic Le Marchand, Christopher A. Haiman, Charleston W.K. Chiang

## Abstract

Polygenic scores (PGS) are promising in stratifying individuals based on the genetic susceptibility to complex diseases or traits. However, the accuracy of PGS models, typically trained in European- or East Asian-ancestry populations, tend to perform poorly in other ethnic minority populations, and their accuracies have not been evaluated for Native Hawaiians. Using body mass index, height, and type-2 diabetes as examples of highly polygenic traits, we evaluated the prediction accuracies of PGS models in a large Native Hawaiian sample from the Multiethnic Cohort with up to 5,300 individuals. We evaluated both publicly available PGS models or genome-wide PGS models trained in this study using the largest available GWAS. We found evidence of lowered prediction accuracies for the PGS models in some cases, particularly for height. We also found that using the Native Hawaiian samples as an optimization cohort during training did not consistently improve PGS performance. Moreover, even the best performing PGS models among Native Hawaiians would have lowered prediction accuracy among the subset of individuals most enriched with Polynesian ancestry. Our findings indicate that factors such as admixture histories, sample size and diversity in GWAS can influence PGS performance for complex traits among Native Hawaiian samples. This study provides an initial survey of PGS performance among Native Hawaiians and exposes the current gaps and challenges associated with improving polygenic prediction models for underrepresented minority populations.

## Introduction

Genome-wide association studies (GWAS) have identified thousands of genetic variants associated with a plethora of human complex traits and diseases [1,2]. The success of GWAS has enabled a burgeoning field of post-GWAS analysis, including the computation of polygenic scores (PGS) for predicting the genetic risk for an individual based on their genotypes and available GWAS summary statistics to common diseases or complex traits [3,4]. On the basis of the GWAS summary statistics, the PGS is calculated as the sum of the trait-associated alleles an individual carries, weighted by the estimated effect sizes of the alleles. Recent developments have focused on extending PGS modeling to include variants and their appropriate weights genome-wide. Such genome-wide PGS tend to be more efficacious in predicting outcome than a simple approach of selecting only genome-wide significantly associated variants [5–7].

As the sample sizes of GWAS increased, the resulting PGS has become more effective in risk predictions, early disease detection, and the development of precision medicine [8,9]. However, available GWAS data are heavily biased towards European-ancestry individuals [10,11], posing a challenge for PGS due to its poor transferability between populations. In particular, PGS trained with European populations exhibit reduced prediction accuracy when applied to non-European populations [12,13]. This poor transferability has been demonstrated for several ethnic minority populations but has not been evaluated for Native Hawaiians, who make up the largest Pacific Islander population in the U.S. [14].

Native Hawaiians are known to have a higher risk of obesity, diabetes, and cardiovascular disease, among others both within Hawai‘i and nationally [15–19]. They also have the highest mortality rate in several types of cancer compared to other ethnic groups [20–22]. In addition, as consequence of centuries of colonization and globalization, Native Hawaiians are largely admixed[23–25], representing ancestry components derived from Polynesian ancestors (∼40%) and recent (within the last 300 years or so) admixtures from European (∼30%) and East Asian (∼29%) immigrants, among others. These ancestries can also be correlated with elevated risks of certain metabolic diseases [15–17,19]. Despite these elevated disease risks, the Native Hawaiian population is largely understudied and underserved [23,25]. We generally lack the genomic resources and knowledge for this population to reap the benefits of genetic research and genomic medicine [23,26].

Predictions based on PGS is one area that is under-investigated for Native Hawaiians and can be potentially improved. To date, no systematic evaluation of PGS, particularly for metabolic traits and diseases such as body mass index (BMI) and type 2 diabetes (T2D), have been conducted for the Native Hawaiians. Given the underrepresentation of Polynesian ancestries in genomic studies and references, poor transferability of PGS models trained in the largest GWAS dataset is expected as is often observed with other populations and ethnic minorities. On the other hand, it is unclear whether the admixture alleviates some of the transferability issues of PGS at the population level. Because admixture levels vary across individuals within a population, even if admixture alleviates some of the transferability issues, it could create disparity within Native Hawaiian communities depending on individual’s genomic similarity to the underrepresented Polynesian ancestries. It is thus crucial to evaluate the transferability of PGS in this population and assess any disparities specific to Native Hawaiians in order to begin bridging this gap.

We conducted the present study using data from the Multiethnic Cohort (MEC) [27]. We leveraged the data of approximately 5,300 Native Hawaiian (MEC-NH), as well as populations as proxies for East Asian- and European ancestries (approximately 19,600 Japanese Americans, MEC-J; approximately 8,500 White Americans, MEC-W), who were genotyped on the Multi-Ethnic Global Array (MEGA) or Global Diversity Array (GDA) arrays to evaluate the prediction accuracy of PGS for BMI, height, and T2D. We trained PGS models using summary statistics from the largest available consortium GWAS from European (EUR), East Asian (EAS), or multi-ethnic populations for BMI, height, and T2D. We focused on these traits as they are the most available and because they are closely linked to obesity, diabetes, and cardiovascular disease – diseases that show elevated risks within the Native Hawaiian population [18,28,29]. Furthermore, T2D was identified as one of the diseases that the Native Hawaiian communities expressed the most concerns [17,30]. Additionally, we assessed the efficacies of published PGS models from the PGS catalog [31] in MEC-NH. In each case, we also investigated the model efficacy in subsets of Native Hawaiians with higher estimated Polynesian ancestry.

We stress that we utilized this study design as a way to examine health disparities within the Native Hawaiian population, and as a way to evaluate how these models may transfer to other Polynesian-ancestry populations. We used empirical (and potentially noisy) estimates of genetic ancestries as a means to assess the impact on the accuracy of currently available PGS models due to admixture over the last 10-12 generations, which is a product of the colonization of the Hawaiian archipelago by Western countries. Interpretations of these estimates beyond the research context are socially complex, and thus should not supplant current practices based on genealogical records and self-reports. These estimates also do not imply any hierarchies or socially meaningful subdivisions in the Native Hawaiian communities.

## Results

### Overview of the study design

Focusing on complex traits and diseases (*i.e.* BMI, height, and T2D) for which large-scale GWAS summary statistics are publicly available, we took two different approaches to construct PGS models for evaluation (**Fig. 1**). We collected from literature the largest East Asian-ancestry (EAS), European-ancestry (EUR), and multi-ancestry (META) meta-analysis GWAS summary statistics for each trait (**Table S1**) to train PGS models. Based on the summary statistics for a particular trait, we used independent MEC subcohorts for as reference for linkage disequilibrium (LD) (N = 500) and for PGS optimization (N = 3000), and then validated the PGS model in an additional held-out sample (N = 1000) (**Methods**). We trained and optimized PGS models in either EUR-, EAS- or multi-ancestry cohorts, and tested the transferability of the best performing genomic PGS models in Native Hawaiians (Design I in **Fig. 1**).

**Fig 1.**
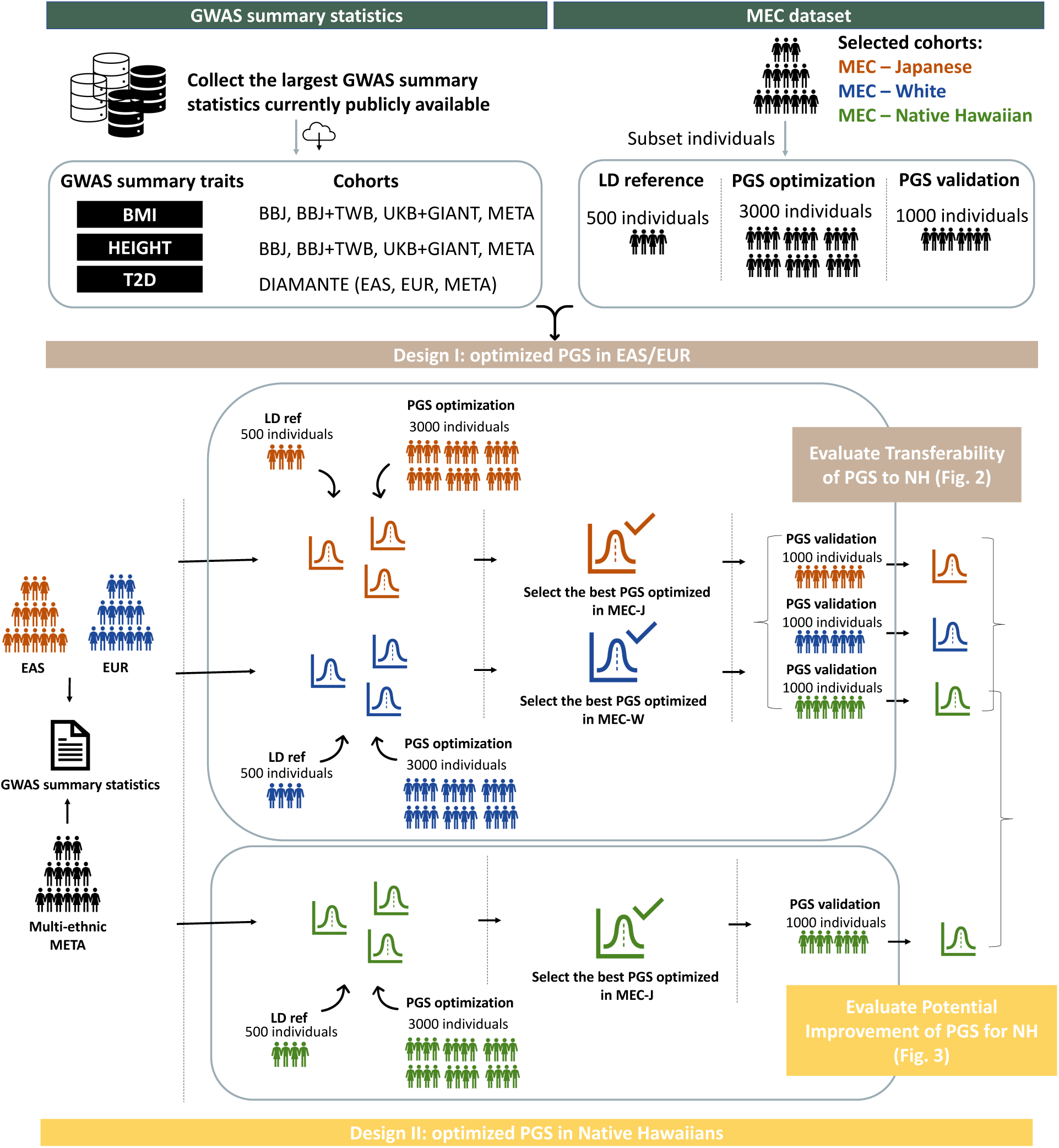
The overall study design of PGS evaluation in Native Hawaiians. The GWAS summary statistics were downloaded from large consortiums and biobanks (BBJ, UKB+GIANT, BBJ+TWB, and META). Each population-specific GWAS (EAS, EUR) were used to train PGS models with the matching MEC cohort as the LD reference and optimization cohort (MEC-J for EAS, MEC-W for EUR). Multi-ancestry meta-analysis GWAS (META) were used to train PGS for both EAS and EUR populations. In Design I, EAS- or EUR-optimized PGS were validated in held-out MEC-J, MEC-W and MEC-NH samples. Comparisons of PGS prediction accuracy between MEC-NH and MEC-J or MEC-W provide the metric for transferability. In Design II, PGS models based on EAS or EUR GWAS were using MEC-NH, and the performance in held-out MEC-NH were then compared to the corresponding metric in Design I to assess potential improvement of prediction by PGS for MEC-NH. See **Table S1** for detailed descriptions of the GWAS datasets used for this study.

Secondly, we explored the potential of using Native Hawaiians as the optimization cohort to improve the accuracies of PGS models within MEC-NH (Design II in **Fig. 1**). Because the Native Hawaiian population, or even the larger Polynesian-ancestry Pacific Islander populations, are generally much smaller in sample sizes compared to the available consortium GWAS from continental populations, it is generally infeasible to amass the sample sizes necessary for a well-powered GWAS that will be informative for PGS construction. We thus investigated whether the MEC-NH could be used for identifying the optimal PGS model to improve upon on the accuracy (hence alleviate the transferability issues) of PGS, even though the GWAS summary statistics were still derived from EAS, EUR, or multi-ancestry meta-analysis.

### Reduced prediction accuracy when applying EAS- or EUR-trained PGS to Native Hawaiians in some scenarios

We first assessed the transferability of PGS to the Native Hawaiian population (Design I in **Fig. 1**). In this case, we identified GWAS summary statistics to build and optimize the PGS model using population-matched cohorts from the MEC, with the Japanese and Whites as representatives of the East Asian and European-ancestry cohorts, respectively (*i.e.* EAS GWAS were optimized using MEC-J; EUR GWAS were optimized using MEC-W; Multi-ancestry GWAS were optimized using either MEC-J or MEC-W separately). We then evaluated the performance of the PGS in held-out MEC-J, MEC-W, and MEC-NH individuals by R^2^. Poor transferability is indicated if there is a noticeable drop-off in prediction accuracy when a PGS model optimized in one population (*e.g.* EAS GWAS optimized in MEC-J) is tested in another population (*e.g.* MEC-W or MEC-NH validation cohort).

As expected, PGS models optimized in MEC-J or MEC-W showed the highest prediction accuracy in validation cohorts from the same populations (**Fig. 2**). For instance, for BMI based on GWAS summary statistics from BBJ, the best PGS model optimized in MEC-J achieved the highest partial R^2^ in held-out MEC-J samples among the validation cohorts tested (partial R^2^ = 0.059), while the best PGS model based on GWAS summary statistics from GIANT+UKB and optimized in MEC-W achieved the highest partial R^2^ in held-out MEC-W (partial R^2^ = 0.088). Moreover, consistent with the expectation of poor transferability, PGS trained in EAS tend to have reduced prediction accuracy in the other continental population. For instance, for BMI, EAS-trained PGS model for BMI had reduced partial R^2^ in MEC-W (0.059 vs. 0.022; one-sided *p* = 0.041 by bootstrapping), and EUR-trained PGS model for BMI performed more poorly in MEC-J (0.088 vs. 0.043; one-sided *p* = 0.045). PGS models trained from multi-ancestry meta-analysis GWAS sometimes reduced the gap in prediction accuracy between MEC-J and MEC-W, though they do not necessarily have higher population-specific prediction accuracies depending on the traits examined (**Fig. 2**).

**Fig 2.**
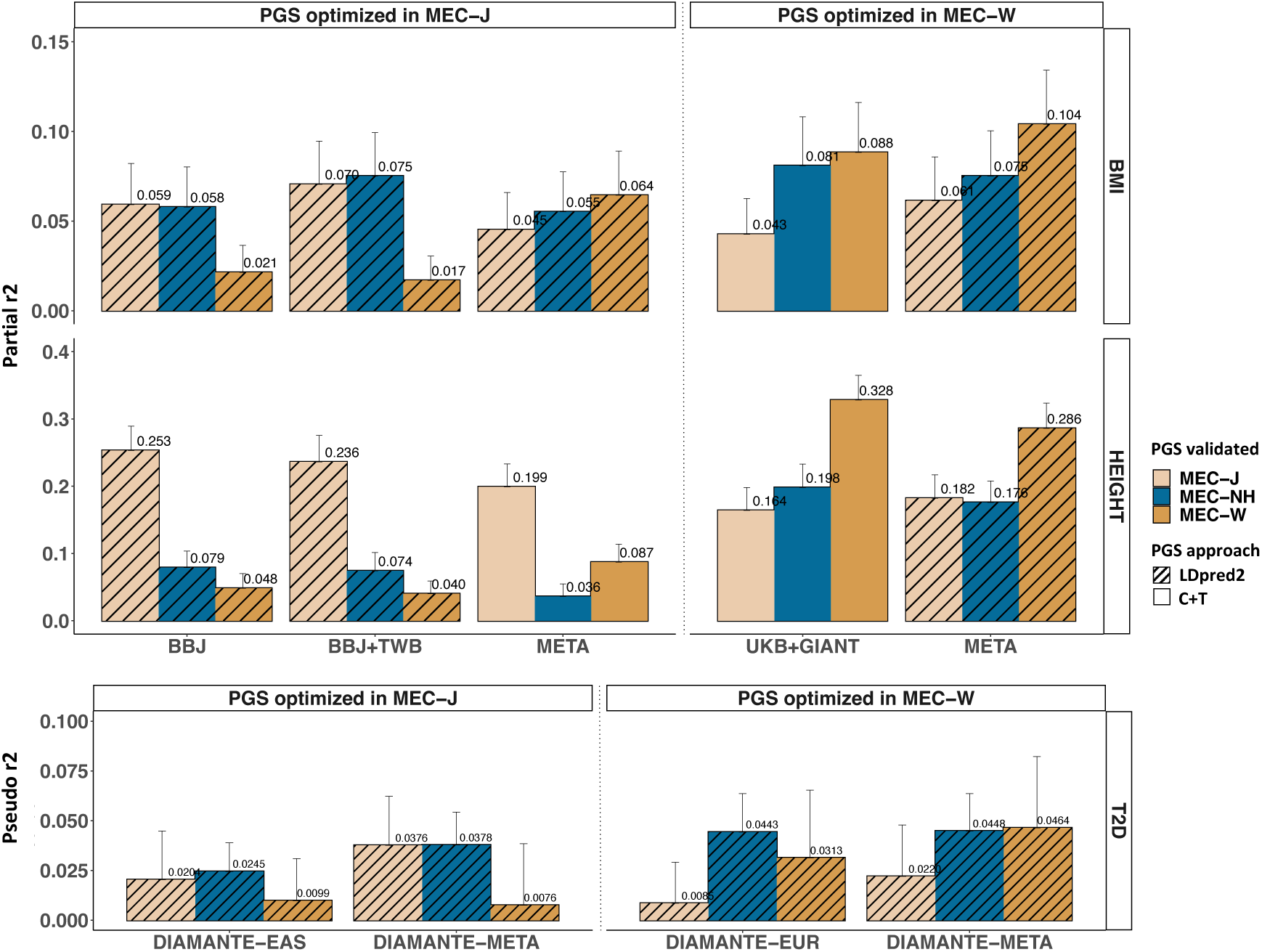
The transferability of EAS- and EUR-trained PGS for BMI, height and T2D. The genomic PGS model with the highest prediction accuracy in optimization cohorts was validated in held-out MEC-J, MEC-W, and MEC-NH cohorts. This figure summarizes the results of analysis in Design I, Fig.1, and details of the model parameter for the best performing PGS model can be found in **Table 1**. Best model based on the C+T or LDpred2 approach are represented by blank or hashed bars, respectively. The standard error for the R^2^ were calculated using 1,000 sets of bootstrap samples. For BMI and height, random 1,000 individuals from each of MEC-J, MEC-W, and MEC-NH were used for validation. For T2D, all cases and controls that were not used in training were used for validation: 3,315 cases and 6,700 controls for MEC-J, 496 cases and 4,063 controls for MEC-W, 392 cases and 549 controls for MEC-NH.

When validating EAS- or EUR-trained PGS in the Native Hawaiian cohort, transferability was not consistently poor, depending on the trait or the GWAS summary statistics used for training (**Fig.2**). For height, there were noticeable reduction in prediction accuracy (*e.g.* for EAS-trained PGS based on the BBJ GWAS, partial R^2^ = 0.253 in MEC-J to 0.048 for MEC-NH; one-sided *p*<0.001). In contrast, EAS- or EUR-trained PGS for BMI showed little drop-off when evaluated in MEC-NH (**Fig.2**). T2D PGS models showed similar pattern as BMI, though the overall prediction accuracies are relatively low (**Fig.2**)

### PGS optimized in Native Hawaiians did not necessarily improve the PGS transferability

In absence of a large-scale powerful GWAS in Polynesian-ancestry populations, another possibility to improve the PGS prediction accuracies for Native Hawaiians may be to use the MEC-NH cohort for optimization, as this may lessen the differences in linkage disequilibrium between the training and validating cohorts and thereby reduce the transferability gap of PGS. Therefore, we employed MEC-NH as an additional optimized cohort, following the same pipeline (Design II, **Fig.1**) to compare with the PGS models optimized using MEC-J and MEC-W. When validated in held-out MEC-NH samples, we observed that generally speaking, PGS models optimized in MEC-NH did not necessarily have better prediction accuracy in the validation cohort (**Fig.3**). NH-optimized PGS models have at best similar prediction accuracies, if not worse (**Fig. 3**). Our results thus suggest that optimization in MEC-NH did not necessarily lead to improved PGS accuracy compared to using the optimization cohort with consistent ancestry as the cohort generated the GWAS summary statistics.

**Fig 3.**
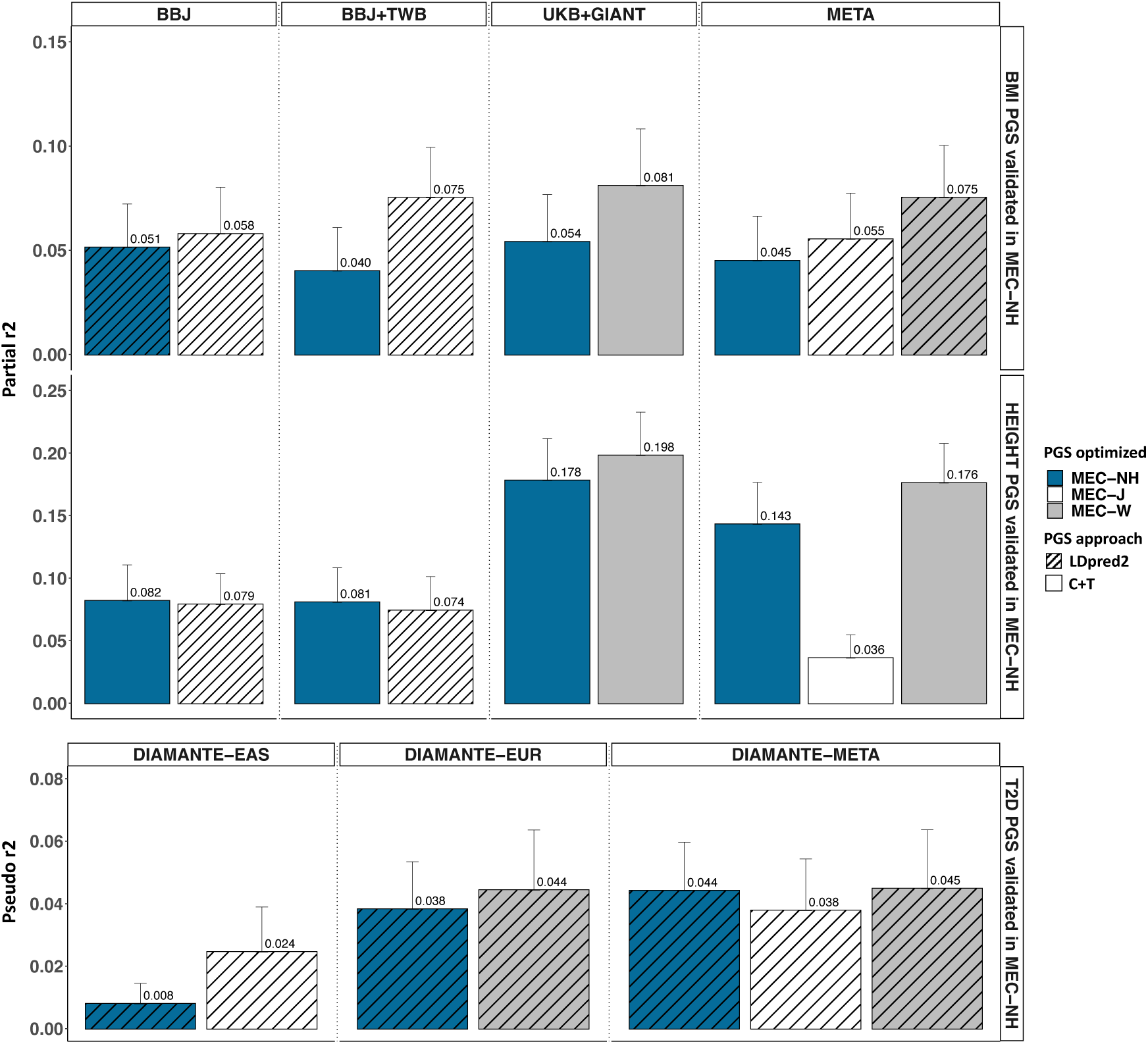
The impact of MEC-NH as optimization cohort on PGS prediction accuracies in held-out MEC-NH for BMI, height, and T2D. For each combination of GWAS-trait PGS models that was previously optimized in MEC-J or MEC-W in Fig.2, the same data was then optimized using MEC-NH samples here (Design II, Fig.1). Previously optimized PGS models and the MEC-NH-optimized models were both validated in the held-out MEC-NH cohort to evaluate if optimization in MEC-NH would improve the prediction accuracy in Native Hawaiians. Details of the model parameter for the best performing PGS model can be found in **Table S2**. Best model based on the C+T or LDpred2 approach are represented by blank or hashed bars, respectively. The standard error for the R^2^ were calculated using 1,000 sets of bootstrap samples. For BMI and height, random 1,000 MEC-NH individuals were used for validation. For T2D, 392 cases and 549 controls from MEC-NH were used for validation.

### Prediction accuracy of publicly available PGS models for Native Hawaiians

We constructed the PGS evaluated here based on limited number of, albeit some of the largest and most recent, GWAS meta-analysis datasets for BMI, height, and T2D (**Table S1**). However, there are a number of published PGS models for these traits in the PGS catalog (**Table S2**, URL https://www.pgscatalog.org/), some of which may have used different GWAS datasets or different methodologies for constructing PGS. We thus validated all of the BMI (N = 11), height (N = 4), and T2D (N = 10) PGS models available as of mid-2022 in MEC-NH, and compared them with our top-performing PGS.

We found that the PGS developed in this study aligns closely with the best-performing public PGS models for BMI and height in terms of prediction accuracy (**Fig. 4**). Moreover, in the case of T2D, our PGS models developed based on the GWAS summary statistics from the DIAMANTE European- or multi-ancestry meta-analysis [32] showed a higher level of predictive accuracy than other PGS models found in the PGS catalog (**Fig.4**).

**Fig 4.**
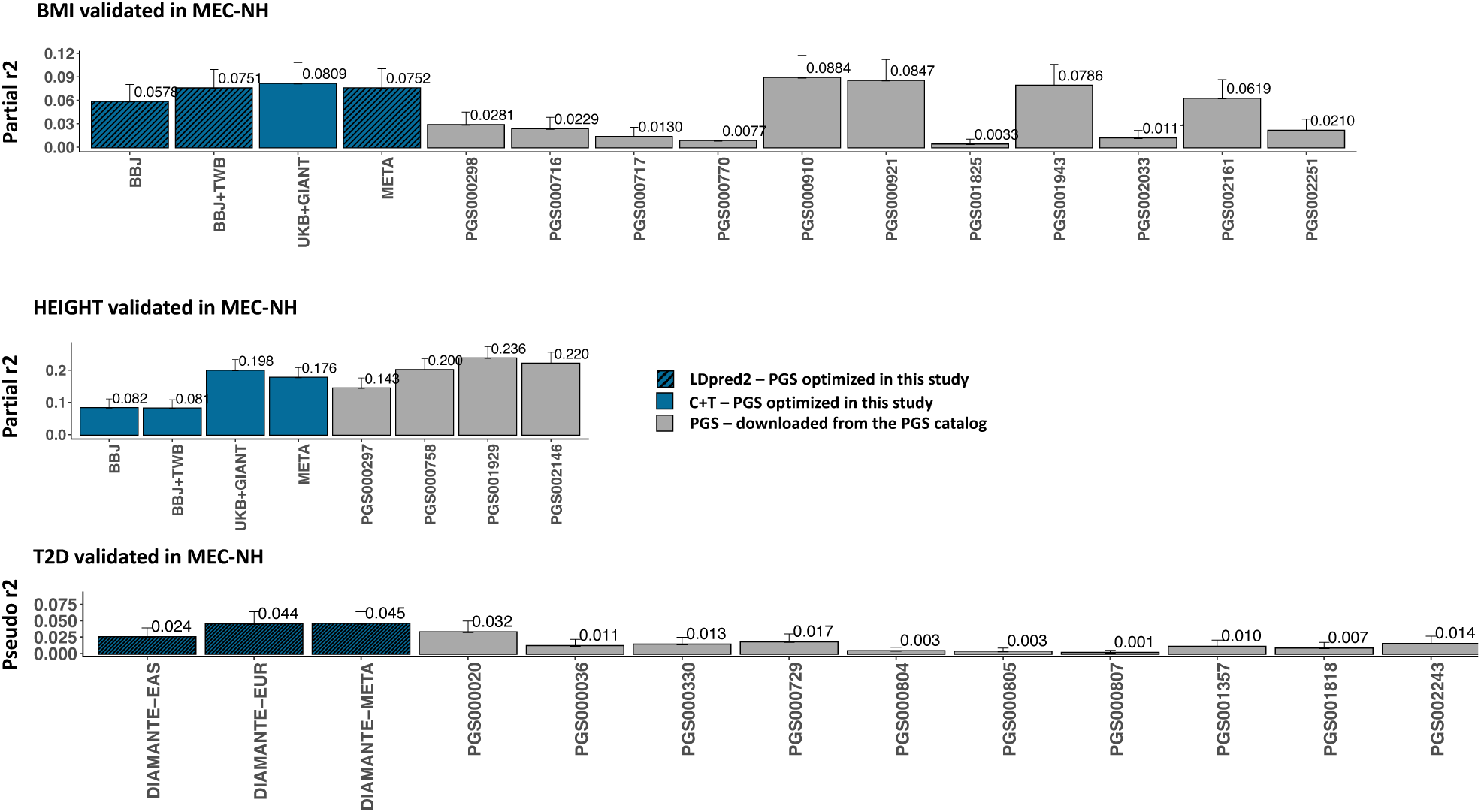
A Comparison of PGS between the optimal PGS from this study and PGS from the PGS-catalog. PGS models available on the PGS catalog (URL https://www.pgscatalog.org/) as of May 18, 2022 were downloaded for BMI, height, and T2D, and validated in the same MEC-NH individuals here. Blue bars represent the PGS constructed in this study with the highest prediction accuracy in MEC-NH. Clear and hash bars indicate the PGS was derived from C+T or LDPred2 approaches. Gray bars depict PGS from the PGS catalog. For BMI and height, random 1,000 MEC-NH individuals were used for validation. For T2D, 392 cases and 549 controls from MEC-NH were used for validation.

### Reduced PGS prediction accuracy in the Native Hawaiians most enriched with Polynesian ancestries

The relatively comparable prediction accuracy of EAS- or EUR-trained PGS models or publicly available PGS models in MEC-NH could be in part driven by admixture. However, Polynesian ancestries exist on a continuum in the population. Because Polynesian ancestries are likely the component of ancestries least similar to the ancestries of individuals participating in the GWAS datasets, there may be disparity in PGS prediction accuracy among individuals with different proportions of Polynesian ancestries [33]. Thus, we explored the accuracy of PGS in subsets of 1000 Native Hawaiians with highest Polynesian ancestries (PNS; minimum estimated proportion of ancestry = 65%), and compared to the 1000 randomly selection individuals. For some PGS models, particularly the best publicly available PGS models for BMI, there was substantially reduced prediction accuracy within the PNS subsets (**Fig.5**). For instance, the partial R^2^ for model PGS00910 was 0.088 in randomly selection MEC-NH, but 0.040 when applied in PNS (one-sided *P =*0.04). In contrast, the PGS model in this study using the MEC-NH for optimization showed relatively little reduction in performance (**Fig. 6**). These findings suggest that while optimizing PGS in MEC-NH may not necessarily lead to improved transferability across Native Hawaiians in general, it could yield enhanced PGS accuracy among Native Hawaiians enriched with Polynesian ancestries, and thus may be more applicable to other Polynesian-ancestry populations across the Pacific. However, because most of the MEC-NH data were used for optimizing the PGS model, we have fewer individuals enriched with Polynesian ancestry to use as validation, resulting in larger error bars and less stable estimates of the prediction accuracy of the PGS models, and preventing us from evaluating PGS models for T2D as we ended with insufficient number of cases and controls.

**Fig 5.**
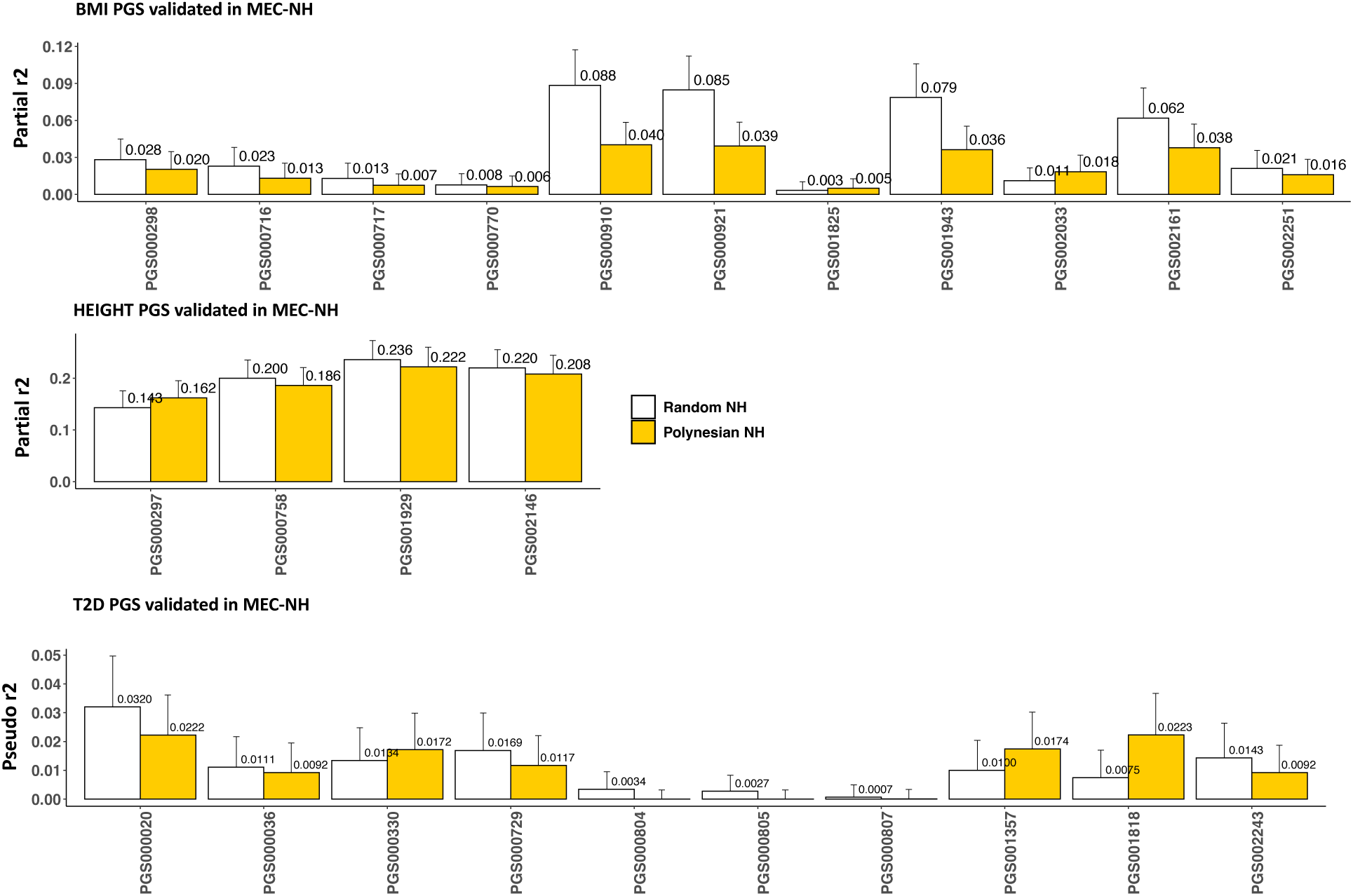
Prediction accuracies of models from PGS catalog in the random Native Hawaiian and Native Hawaiian with highest Polynesian ancestry validation sets. Each PGS model from PGS catalog was assessed in validation datasets from MEC-NH. For BMI and height, the validation cohort consisted of randomly selected 1000 MEC-NH or the 1000 individuals with the highest estimated Polynesian ancestry (minimum estimated Polynesian ancestry = 65%) among the entire MEC-NH cohort. This was not restricted to the 1,000 individuals reserved for validation in Figures 2-4 as none of the MEC-NH individuals were used in construction of the publicly available PGS models. For T2D, because only 768 individuals (346 cases, 422 controls) with > 65% estimated Polynesian ancestry were available, we compared to 768 randomly selected MEC-NH individuals (318 cases, 450 controls).

**Fig 6.**
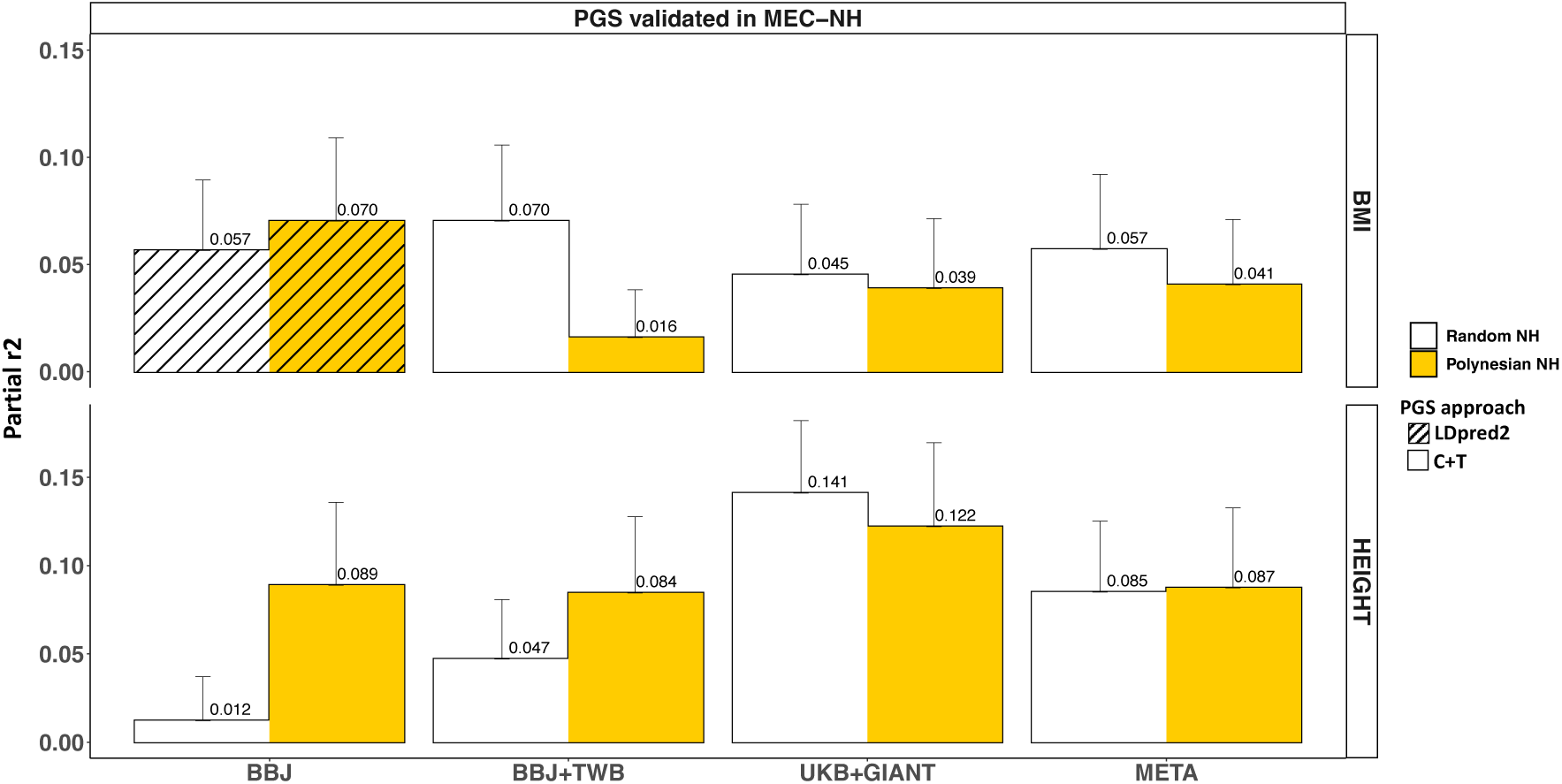
Prediction accuracies of models from this study in the random Native Hawaiian and Native Hawaiian with highest Polynesian ancestry validation sets. The PGS models were all trained from EAS-, EUR- or multi-ancestry GWAS, but using the MEC-NH for optimization. The resulting models were then validated in 200 randomly selected MEC-NH individuals and the same number of individuals with highest estimated Polynesian ancestries (minimum estimated Polynesian ancestry = 65%). We did not perform the analysis for T2D due to too few case / control samples, particularly among those with high Polynesian ancestries.

## Discussion

In this study we conducted a systematic assessment of the prediction accuracies and transferability of PGS models for Native Hawaiians. We assessed either PGS models constructed in this study or models that are publicly available from the PGS catalog. We focused on BMI, height, and T2D, as these are traits or diseases that showed different distributions between Native Hawaiians and other continental populations, and that Polynesian ancestries may be correlated with disease risk [15]. While we observed that EAS-trained PGS models have reduced prediction accuracy in MEC-W (the MEC cohort representing EUR ancestries) and vice versa, our results revealed that these PGS models at times showed comparable prediction accuracies in the Native Hawaiian cohort, especially for BMI and T2D. Empowered by the Native Hawaiian cohort in the Multiethnic Cohort Study (N ∼ 5,300 with genome-wide genotyping array data), we were also able to evaluate whether using the MEC-NH individuals for optimizing the PGS model could improve PGS prediction accuracies for this population, even though our findings suggested limited improvements. Furthermore, because the Native Hawaiian population displays a continuum of genetic ancestry due to its colonial history, with Polynesian ancestries being the majority ancestry, we also evaluated PGS prediction accuracies as function of this ancestry. For publicly available PGS models for BMI and T2D, we did observe a reduction in prediction accuracy when applied to the Native Hawaiians most enriched with Polynesian ancestries. Our results suggest that recent admixture with European or East Asian ancestries are mediating the PGS performance, and these PGS models in general may be even less applicable to the Polynesian-ancestry populations across the Pacifics at large.

Across the traits, populations, and GWAS datasets that we examined here for training PGS models for Native Hawaiians, irrespective of the degree of reduction in prediction accuracy, we found that training with European-based GWA S generally produced better performing PGS models. For instance, the prediction accuracy by partial R^2^ of height PGS model from the UKB+GIANT dataset (N = ∼700,000), optimized in MEC-W, was 0.198 in validation MEC-NH, compared to 0.074 for PGS model derived from BBJ+TWB GWAS data (N = 102,900) with optimization in MEC-J (one-sided *P* < 0.001; **Fig.2**). This is thus reflective of the bias in GWAS, where ancestry-specific GWAS for European-ancestry populations are still much larger, and thus more powered and informative for PGS, than East Asian-ancestry populations. Another hint of this bias can also be observed when PGS was constructed from multi-ancestry meta-analysis data. In this case, the same GWAS dataset was optimized in MEC-W or MEC-J separately, but the resulting PGS still had higher prediction accuracy in held-out MEC-W (partial R^2^ = 0.286) than MEC-J (0.199; one-sided *P =* 0.023), respectively. This is potentially due to the European-ancestry populations still representing the majority of the dataset in the multi-ancestry meta-analysis (∼628,000 European-ancestry individuals vs. ∼ 179,000 East Asian-ancestry individuals in Sakaue S., *et al*. [34]). Therefore, in addition to increasing sample sizes of future GWAS, increasing representation and diversity of these GWAS will also help improve the PGS accuracy, particularly for a population such as the Native Hawaiians (or African Americans and Latinos) who are not a major constituent of multi-ancestry GWAS yet. This may be particularly important since using the diverse population for optimization, without fundamentally changing the representation of the GWAS data, did not appear to make meaningful improvements (**Fig.3**).

While we generally advocate for greater inclusion of diverse populations in future GWAS, which both increase the sample size and the diversity of the GWAS, it could also be difficult to predict the impact on prediction accuracy of the PGS and may be dependent on the trait architecture. For instance, the same multi-ancestry meta-analysis cohort optimized in MEC-W individuals showed an improvement in prediction accuracy for BMI (partial R^2^ = 0.104, compared to partial R^2^ = 0.088 in UKB+GIANT only; *P* < 0.001 by paired bootstrapping; **Fig.2**), but a reduction for height (partial R^2^ = 0.286 in multi-ancestry meta-analysis, compared to R^2^ = 0.328 in UKB+GIANT; *P* < 0.001; **Fig.2**). For Native Hawaiians, switching from a smaller European-only GWAS to a marginally larger multi-ancestry GWAS did not improve the prediction accuracies for either trait, though the performance was comparable (partial R^2^ = 0.081 vs. 0.078 for BMI, 0.198 vs. 0.176 for height; **Fig. 2**). It is therefore important to assess the prediction accuracy empirically, particularly for the diverse ethnic minority populations such as the Native Hawaiians.

In addition to sample size and diversity of the GWAS dataset for training, the size of the optimization and validation samples may also play a role in determining the transferability of the resulting PGS model. In particular, we had compared the PGS models constructed here with those publicly available on the PGS catalog (**Fig. 4**). For some traits, our models may have used larger GWAS but achieved only comparable, if not lower, prediction accuracies compared to the published PGS models. For instance, in the case of height PGS, PGS001929 [35] showed significantly better performance than our best model (partial R2 = 0.236 vs. 0.198; *P =* 0.02 ; **Fig.4**). PGS001929 utilizes only the UKB GWAS (N ∼500,000) and employs the same LDpred2 for PGS construction. The improved performance in PGS001929 may be attributed to a larger optimized dataset and validation sample sizes (N = 349,991 and 43,631, respectively). Our observations here then suggest that the most effective study design to construct PGS for underrepresented admixed populations may be using the largest and most diverse GWAS along with a large sample for optimization. This is yet another challenge for underrepresented minority populations such as the Native Hawaiians when sample sizes are generally small.

In summary, our study informs on PGS performance and transferability in the Native Hawaiians and individuals with Polynesian ancestry. We found that continental-level admixture, over the last 300 years or so, may have mediated the PGS performance for these complex traits in Native Hawaiians. We again stress that our use to quantify proportion of ancestries was solely for the purpose of research use, as a means to evaluate potential disparity in the way PGS can be deployed within the Native Hawaiian population. These estimates are potentially noisy, dependent on external references used, and could induce social harm when taken out of the particular research context. In fact, one future approach to evaluate PGS performance may focus on individual-level metric, where one’s genetic information is evaluated in context of genetic similarity to, say, the European cohort underlying the GWAS study[33], rather than explicit quantification of proportion from a labeled ancestry. We also found that sample size and diversity in GWAS, as well as the availability of large optimization cohort, may complicate the construction and application of PGS in underrepresented populations. This is another obstacle for Native Hawaiians, as collection and use of genetic data from indigenous populations are fraught with past misuse that led to general mistrust from the community[36–38]. For a population already small in size, the availability of cohorts to optimize these PGS models are also much smaller than that available for other continental populations, which in turn harms the community in reaping the benefits of genomic medicine. Thus, an ongoing discussion, engagement, and involvement of the community centered around issues of participation in research, genetic data collection, and data sovereignty will be required. Finally, future studies may also focus on traits and diseases that we did not study here, such as cardiovascular diseases that the Native Hawaiians are also known to be susceptible [18,39], as well as traits that the Native Hawaiian communities expressed explicit concern and should be made as research focus, such as asthma [17,30], so to further encourage community participation in research. It is with the multi-pronged strategy of inclusion in genetics research, reduction of biases, and focus on community concerns that we can reduce the disparity in PGS model performance in this case, and alleviate health disparity in general.

## Material and Methods

### GWAS summary statistics and PGS Catalog datasets

To comprehensively evaluate the transferability of PGS for the Native Hawaiian population, an admixed population with substantial East Asian (EAS) and European (EUR) ancestries [19], we have utilized EAS and EUR GWAS, as well as multi-ethnic GWAS, summary statistics for training PGS models for BMI, height, and T2D. We collected and curated the largest available GWAS summary statistics data from the most recent publications [32,34,40–43] (**Table S1**). In addition to training PGS models from these latest GWAS, we also obtained and tested all models from the PGS-catalog database (http://www.pgscatalog.org/) related to these three traits, published in the database as of 05/18/2022 (**Table S4**).

In all cases, we downloaded the datasets in GRCh38 coordinates or converted the coordinates to GRCh38 genome build using triple LiftOver [44] to ensure alleles are aligned in genomic regions that may have inverted between genome builds. Additionally, we verified the order of reference and alternative alleles in the GWAS summary data by comparing it to the GRCh38 reference genome. We also removed the indels in the GWAS summary statistics data.

### Study cohorts and quality controls of individual data

To optimize the PGS models for each trait, and to evaluate the prediction accuracy and transferability of each PGS model, we leveraged the individual genetic data from the Multiethnic Cohort (MEC) study [27]. MEC encompasses five major ethnic groups: Japanese Americans, Native Hawaiians, African Americans, Latinos, and Non-Latino Whites, with up to approximately 70,000 individuals with genome-wide array data. We utilized the subcohorts genotyped with the Illumina Multi-Ethnic Global Array (MEGA) and Global Diversity Array (GDA) arrays, in total containing 19,677 (MEGA: 5,022; GDA: 14,655) Japanese Americans (MEC-J), 11,316 (MEGA: 829; GDA: 10,487) Non-Latino White (MEC-W), and 5,388 (MEGA: 4,144; GDA: 1,244) Native Hawaiians (MEC-NH). MEC-J and MEC-W subcohorts were used as benchmarks for the prediction accuracies of PGS trained and optimized in East Asian and European populations, respectively.

We removed population outliers for each population using the results from unsupervised ADMIXTURE v1.3.0 [45] with the 1000 Genomes Project populations to guide the interpretation of inferred ancestry components. We pruned the SNPs for linkage disequilibrium (LD) using the --geno 0.05 and --indep-pairse 100 10 0.1 commands in PLINK v1.9[46]. For the ADMIXTURE analysis, we set K=5, and each K was calculated for 5 repetitions. For MEC-J and MEC-W, we removed individuals admixed within the last 1-2 generations and defined a relatively unadmixed subset based on an inferred minimum of 95% genetic components of East Asian and European ancestries, respectively. While fully recognizing that genetic ancestries exist on a continuum, and any attempt to define a discrete population on the basis of inferred genetic ancestry will be arbitrary, we opted to rely on inferred genetic ancestry to define our EAS and EUR populations in alignment with the common practices in the field and with the ancestries of the source populations for the GWAS data we used. In total, we retained 18,705 individuals in MEC-J (MEGA: 4,828; GDA: 13,877) and 8,532 individuals in MEC-W (MEGA: 634; GDA: 7,898) for analysis. We did not filter out any self-reported Native Hawaiian individuals on the basis of estimated genetic ancestry, since it is a well-known admixed population with a continuous cline of any major ancestry components that can be estimated [15]. Furthermore, it is the community belief that estimated genetic ancestries should not be used as an exclusion criterion for community membership and should not supplant well-established custom of self-identity or genealogical records. Therefore, we included all self-reported Native Hawaiians within the MEC as a single population, although at times we do evaluate the efficacy of PGS models for subset of MEC-NH with higher proportion of estimated Polynesian ancestry, both for a better evaluation of using PGS in realizing precision medicine at the individual level and for assessment of the generalizability of PGS models to other Polynesian-ancestry populations.

To avoid overfitting PGS models in validation cohort, we randomly subset the individuals into three non-overlapping groups when evaluating PGS models for BMI and height: 500 individuals per population for LD reference, 3000 individuals per population for optimizing the PGS model, and 1000 individuals per population for validating the PGS model (**Fig. 1**). For T2D, we randomly selected 800 cases as well as 1500 controls for a total of 2300 individuals per population for optimization. We then used all remaining cases and controls from each cohort for validation: 3315 cases and 6700 controls for MEC-J, 496 cases and 4063 controls for MEC-W, 392 cases and 549 controls for MEC-NH. T2D cases were defined based on any self-reported T2D diagnosis by a physician or medical professional, or the use of medication for treatment of diabetes, on questionnaire 1 to 5 of the MEC survey or based on Medicare claim data based on ICD-9 codes (249-250.99) or ICD-10 codes (E11.X [47,48]). using information from the phenotype file, including patient age and T2D status. This process involves tracking individuals over time, considering records across decades. If an individual’s T2D status changes from negative to positive in later traces, the record is updated accordingly. Our strategy entails evaluating all available records for each person across five time points. Records with missing data are excluded, ensuring the reliability of our findings. For individuals used as LD reference or as validation, we randomly selected individuals who were genotyped on the MEGA array or GDA array, respectively, for each of MEC-J, MEC-W, and MEC-NH. For individuals used in optimizing PGS model, we selected from those genotyped on the MEGA array for MEC-J and MEC-NH and from those genotyped on the GDA array for MEC-W due to availability of samples.

### Polygenic Score development and validation

Two commonly used approaches were utilized in this study to construct the PGS model: Clumping and Thresholding (C+T) [6,49] and LDpred2 [7]. The C+T method involved clumping SNPs in LD based on r^2^, and distance (kb) parameters, and thresholding SNPs based on specific thresholds of p-values [50–52]. On the other hand, LDpred2 employed the Bayesian method to estimate the effects of genetic variants on a specific trait and considered the LD information between genetic variants [53]. We followed previous studies [54] in constructing PGS models from C+T and LDpred2, with the following modifications. For the C+T, we used p-values thresholds of: 0.1, 0.2, 0.5, 0.05, 0.01, 0.005, 1e-3, 5e-4, 1e-4, 5e-5, 1e-5, 5e-6, 1e-6, 5e-7, 1e-7, 5e-8; r2 values (based on the LD reference sample of 500 individuals) of: 0.2, 0.1, 0.01, 0.005, and distance in kilobases (kb) window sizes: 250, 500. Together we evaluated 128 combinations of parameters to identify the optimal PGS (by partial R^2^ and pseudo R^2^ for quantitative and dichotomous traits, respectively; see below) in the optimization sample of 3000 individuals. For LDpred2, we used a grid of values for hyper-parameters/tuning parameter - causal variants (ρ), ℎ^2^ (the SNP heritability), and sparsity (whether to fit some variant effects to exactly zero) to construct PGS. We used ρ from a sequence of 17 values from 0.01 to 1 on a log-scale, a range of ℎ^2^ within (0.7, 1, 1.4) × estimated heritability, and a binary sparsity option of either on and off (LDPred2-grid models). In addition, we tested a model assuming infinitesimal causal effects, where each variant assumed to contribute to disease risk (LDPred2-inf model). In total, we evaluated 103 PRS models using LDPred2.

To evaluate the association of a PGS model in either the optimizing sample or validation sampling, we evaluated a regression model using the PGS for each individual as the predictor variable and the trait or disease as response variable using R (version 4.0.0) [55]. For BMI and height, the trait was stratified by sex, and regressed against age and age^2^. We extracted the residuals from the model using the R package stats v3.6.2 [55] and the residuals() function, and inverse normalized the residuals using the R package norm v1.0 [55] and the qnorm() function. Finally, we merged the results from males and females. For covariates, we additionally included ten Principal Component Analysis (PCA) data to adjust for the impact due to ancestry and population structure. For T2D, we used logistic regression, with covariates age, sex, and ten principal components. The primary metric of PGS model efficacy is the partial R^2^ for quantitative traits and pseudo R^2^ for dichotomous traits. Partial R^2^ was calculated using the R package rsq v2.5 [56] and its function rsq.partial(). Pseudo R^2^ was calculated using the R package DescTools v0.99.43 [57] and its function PseudoR2(). To obtain the pseudo R^2^ with only the PGS score effect in the dichotomous trait, we calculate the difference in pseudo R^2^ between the full model (including PGS) and the partial model (without the PGS score). To obtain confidence interval for the partial or pseudo R^2^, we computed the standard error from 1000 bootstrap samples. Transferability of PGS is evaluated based on differences of the partial or pseudo R^2^ between two populations or between two PGS models. Significance is calculated based on testing differences in mean of the partial or pseudo R^2^ distribution from the bootstrap samples.

## Data Availability

Genotype data is available on dbGaP with accession numbers phs 000220.v2.p2 and phs002183.v1.p1

## Data and code availability

Genetic data utilized in this study will be available on dbGAP (accession number: phs002183.v1.p1). GWAS summary statistics were downloaded from literature, see **Table S3** for reference. For the downloaded PGS models from the PGS catalog, see **Table S4**. We built a package to incorporate the pipeline for training, optimizing, and validating the PGS models. The package is publicly available on GitHub: https://github.com/imyingchulo/gprs.

## Supporting information

**Table S1. Parameters of the best genomic PGS models in Design I.**

For each combination of complex trait, GWAS dataset, and optimization cohort, this table lists the model parameters and performance (in the optimization cohort) for the best genomic PGS model. To optimize, each combination of trait and GWAS was evaluated in a grid-like search in the optimization cohort, with the best performing model among the candidate 128 C+T models and 103 LDpred2 models. Across all combinations of GWAS dataset and traits, LDpred2 produced the best model based on partial R^2^ (for BMI and height) or pseudo R^2^ (for T2D) in the optimization cohort in 11 out of 14 instances.

**Table S2. Parameters of optimal genomic PGS models when optimized in MEC-NH (Design II).** As is the case for Table S1, but the optimization cohort is in MEC-NH in each case.

**Table S3. Details of GWAS summary statistics and references used in this study.**

This table provides the overview of GWAS datasets used in this study, including the data resources, cohort, SNPs, genome built, and population size.

**Table S4. Additional information of PGS models from PGS catalog evaluated in this study** This table provides the references and additional information for the 39 PGS models included in this study.

## Acknowledgments

We would like to thank David Conti and Hailiang Huang for discussions of the analysis and release of pre-publication summary statistics. We would also like to thank the Native Hawaiian participants in the Multiethnic Cohort that are involved in this study. The Multiethnic Cohort was funded through grants from the National Cancer Institute (U01CA164973, P01CA168530) and National Human Genome Research Institute (U01HG007397). We also would like to thank University of Hawai‘i Cancer Center’s Native Hawaiian Community Advisory Board for reviewing the study proposal and providing comments to earlier versions of this manuscript. This study is supported by grants from the National Human Genome Research Institute (R01HG011646 to C.W.K.C.) and the Taiwan-USC Postdoctoral Fellowship from Taiwan Education Ministry. Computation for this work was supported by the University of Southern California’s Center for High-Performance Computing (https://hpcc.usc.edu).

## Notes

### Competing Interest Statement

The authors have declared no competing interest.

### Author Declarations

Genetic data utilized in this study will be available on dbGAP (accession number: phs002183.v1.p1). GWAS summary statistics were downloaded from literature. For the downloaded PGS models were from the PGS catalog. We built a package to incorporate the pipeline for training, optimizing, and validating the PGS models. The package is publicly available on GitHub: https://github.com/imyingchulo/gprs.

